# Genetic characterization of the ALFA study: Uncovering genetic profiles in the Alzheimer’s *continuum*

**DOI:** 10.1101/2023.04.26.23289138

**Authors:** Natalia Vilor-Tejedor, Patricia Genius, Blanca Rodríguez-Fernández, Carolina Minguillón, Iman Sadeghi, Armand González-Escalante, Marta Crous-Bou, Marc Suárez-Calvet, Oriol Grau-Rivera, Anna Brugulat-Serrat, Gonzalo Sanchez-Benavides, Manel Esteller, Karine Fauria, José Luis Molinuevo, Arcadi Navarro, Juan Domingo Gispert, the Alzheimer’s Disease Neuroimaging Initiative, the ALFA study

**Affiliations:** Barcelonaβeta Brain Research Center (BBRC), Pasqual Maragall Foundation, Barcelona, Spain; Centre for Genomic Regulation (CRG), The Barcelona Institute for Science and Technology, Barcelona, Spain; Department of Clinical Genetics, Erasmus University Medical Center, Rotterdam, Netherlands; Universitat Pompeu Fabra, Barcelona, Spain; IMIM - Hospital del Mar Medical Research Institute. Barcelona, Spain; Centro de Investigación Biomédica en Red de Fragilidad y Envejecimiento Saludable (CIBER-FES). Instituto de Salud Carlos III, Madrid, Spain; Department of Epidemiology, Harvard T.H. Chan School of Public Health. Boston, MA, USA; Catalan Institute of Oncology (ICO)-Bellvitge Biomedical Research Center (IDIBELL), Hospitalet del Llobregat, Spain; Servei de Neurologia, Hospital del Mar, Barcelona, Spain; Global Brain Health Institute, San Francisco, CA, USA; Josep Carreras Leukaemia Research Institute (IJC), Badalona, Barcelona, Spain; Centro de Investigación Biomédica en Red Cáncer (CIBERONC), Madrid, Spain; Integrated Pharmacology and Systems Neurosciences, IMIM-Hospital del Mar Medical Research Institute, Barcelona, Spain; Institució Catalana de Recerca i Estudis Avançats (ICREA), Barcelona, Catalonia, Spain; Physiological Sciences Department, School of Medicine and Health Sciences, University of Barcelona (UB), Barcelona, Catalonia, Spain; H. Lundbeck A/S, Copenhagen, Denmark; Institute of Evolutionary Biology (CSIC-UPF), Department of Experimental and Health Sciences, Universitat Pompeu Fabra; Centro de Investigación Biomédica en Red Bioingeniería, Biomateriales y Nanomedicina. Instituto de Salud carlos III, Madrid, Spain

**Keywords:** AD *continuum*, ALFA study, Alzheimer’s disease, Neurogenetics, Neurological diseases, Prevention

## Abstract

In 2013, the ALFA (ALzheimer and FAmilies) project was established to investigate pathophysiological changes in preclinical Alzheimer’s disease (AD), and to foster research on early detection and preventive interventions. Since then, it has prospectively followed cognitively unimpaired late/middle-aged participants, most of whom are adult children of AD patients. Risk stratification of cognitively unimpaired individuals, including genetic factors is key for implementing AD prevention strategies. Here, we report the genetic characterization of ALFA participants with respect to neurodegenerative/cerebrovascular diseases, AD biomarkers, brain endophenotypes, risk factors and aging biomarkers, emphasizing amyloid/tau status and gender differences. We additionally compared AD risk in ALFA to that across the full disease spectrum from the Alzheimer’s Disease Neuroimaging Initiative (ADNI). Results show that the ALFA project has been successful at establishing a cohort of cognitively unimpaired individuals at high genetic risk of AD. It is, therefore, well-suited to study early pathophysiological changes in the preclinical AD *continuum*.

**Highlights:**

- Prevalence of ε4 carriers in ALFA is higher than in the general European population.
- The ALFA study is highly enriched in AD genetic risk factors beyond *APOE*.
- AD genetic profiles in ALFA are similar to clinical groups along the *continuum*.
- ALFA has succeeded in establishing a cohort of CU individuals at high genetic AD risk.
- ALFA is well suited to study pathogenic events/early pathophysiological changes in AD.

## 1. Background: Initial progress in genetic research within the ALFA study

In 2013, the ALzheimer’s and FAmilies (ALFA) project was launched by the Barcelonaβeta Brain Research Center with the aim of enhancing our understanding of the pathogenesis and pathophysiology of Alzheimer’s disease (AD) at its early preclinical stages (Molinuevo et al., 2016). The ALFA project consists of the ALFA parent cohort (NCT01835717), and the nested ALFA+ cohort study (NCT02485730). The ALFA parent cohort is composed of 2,743 cognitively unimpaired participants aged between 45 and 74 years at the time of recruitment who underwent cognitive testing, clinical history, magnetic resonance imaging (MRI), environmental and lifestyle questionnaires, and blood sampling. A significant feature of ALFA is that it is a genetically enriched cohort of cognitively unimpaired individuals, as ∼50% of them are adult children of patients with AD dementia diagnosed before the age of 75 years. The recruitment strategy of the ALFA parent cohort has resulted in a population that is enriched in genetic risk factors for AD, the most notable of which is the *ApolipoproteinE* (*APOE*)-ε*4* allele. A subset of the ALFA parent cohort participants were invited to participate in a nested longitudinal long-term study, which is referred to as ALFA+. This study involved more detailed phenotyping, including fluid (blood and cerebrospinal fluid [CSF]) and Positron Emission Tomography (PET). ALFA+ participants were selected according to their age, sex and *APOE-*ε*4* carriership to cover the full range of the AD risk spectrum.

In a pilot study (NCT02198586), ∼575 ALFA parent cohort participants underwent MRI to investigate the effects of *APOE* variants on different cerebral phenotypes. As a result of this project, we found that ε*4*-homozygotes displayed an age-related increase in radial but not axial diffusivity, consistent with a reduced myelin sheath in several white matter regions (Operto et al., 2018, 2019). When assessing gray matter volumes in *APOE*-ε*4* homozygotes, we found reductions in brain regions known to undergo atrophy in symptomatic AD stages (Cacciaglia et al., 2018) and a dose-dependent protective effects of the *APOE*-ε*2* allele (Salvadó, et al., 2021). We also showed, as suggested in previous studies, that *APOE*-ε*4* reversed the association between cognitive performance and brain morphology, similar to aging, suggesting that this risk allele leads to an accelerated biological phenotype of brain aging (Cacciaglia et al., 2019). Furthermore, ε*4*-homozygotes displayed reduced connectivity between networks in areas typically susceptible to amyloid deposition in the early AD *continuum*, and altered effects of amyloid on brain structure (Cacciaglia et al., 2020, 2022). Additional results indicated that cognitively unimpaired *APOE*-ε*4* homozygotes were at significantly higher risk of having pathological levels of white matter hyperintensities (WHM) than heterozygotes (Rojas et al., 2018), and observed a protective effect of the ε*2 allele* on global WMH (Salvadó et al., 2019). We also found a higher prevalence of cerebral microbleeds in *APOE*-ε*4* carriers (Ingala et al., 2020), although no association was found between *APOE-*ε*4* and the enlargement of perivascular spaces (Vilor-Tejedor et al., 2021).

These findings have encouraged us to extend the genetic characterization of the ALFA parent cohort beyond *APOE* to deepen our understanding of the biological mechanisms associated with genetic predisposition to AD at preclinical stages, which may aid in the implementation of prevention programs. In this article, we present the rationale, methods, and genetic characterization of participants of the ALFA parent cohort. In addition, we assessed AD genetic risk prediction across the spectrum of the disease by including data from the Alzheimer’s Disease Neuroimaging Initiative (ADNI) project (Weber, et al., 2021).

## 2. Methods

### 2.1. Design

The 17th September 2012, a press conference was held where the main aims of the ALFA project were presented along with detailed inclusion and exclusion criteria (Molinuevo et al., 2016). Briefly, participants had to be cognitively unimpaired Spanish and/or Catalan-speaking individuals aged 45-74 who agreed to undergo clinical interviews and questionnaires associated with dementia risk factors, cognitive tests, a blood sample extraction for deoxyribonucleic acid (DNA) analysis, and MRI. A total of 2,743 individuals were recruited in the ALFA parent cohort, with 86.3% reporting a diagnosis of AD in at least one of their parents. When considering a more strict family history encoding, 47.4% of the ALFA study participants had at least one of their parents that had been diagnosed with AD before the age of 75 years. In total, 2,686 participants were genotyped, as 57 individuals were excluded since blood extraction could not be performed or sufficient DNA could not be obtained to perform the genotyping. The genetic data processing procedure is detailed in **Supplementary Figure 1**. Summary information on project participants [**Figure 1**], DNA extraction, genotyping, and data availability is described in **Supplementary methods**. A subset of ALFA parent cohort (N=380), referred to as ALFA+, also underwent lumbar puncture to determine biomarkers in CSF, enhanced MRI, more detailed cognitive testing, blood sampling for biomarker determination, ^18^F-fluoro-2-deoxyglucose PET, amyloid, and tau PET imaging. Participants were classified into groups defined by their biomarker profile according to the A/T framework (Jack et al., 2016). CSF amyloid-β 42 (Aβ42) levels of ≤ 1098 pg/ml were designated as A+, and phosphorylated tau (pTau) levels of ≥ 24 pg/ml were designated as T+ (Milà-Alomà et al., 2020).

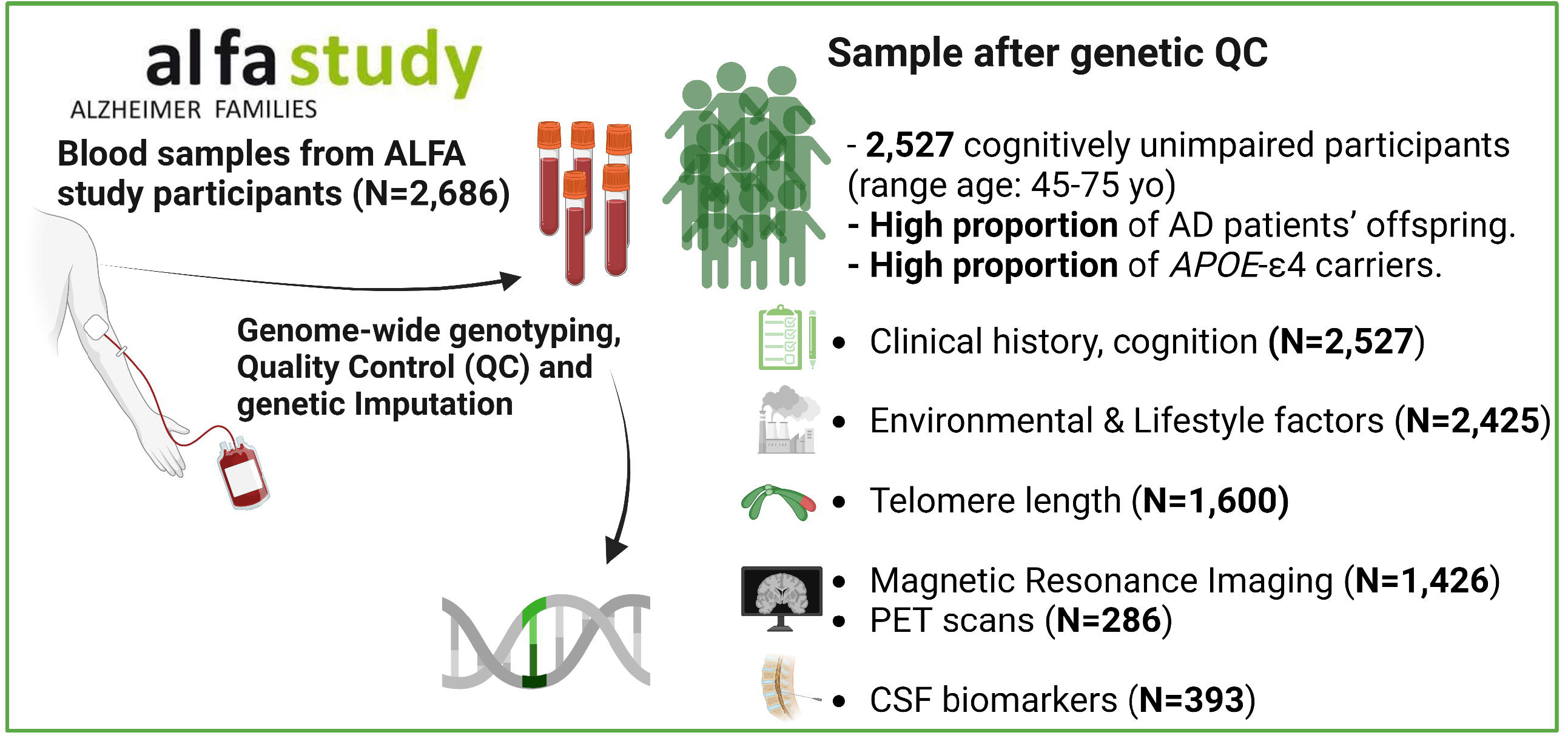

### 2.2. Genetic Quality Control and Imputation

Quality control (QC) of genetic data was performed with the PLINK software. Samples with a call rate of less than 98%, mismatched genetically determined sex (from X-chromosome heterogeneity) *versus* that coming from demographic data, or excess of heterozygosity (4 standard deviations from the mean) estimated by an F statistic, were excluded from the analyses. Individuals at high genetic relatedness (at the level of cousins or closer relatives), sharing proportionally more than 18.5% of alleles (IBD > 0.185), were also excluded. After completion of sample level QC, genetic variants with low minor allele frequency (MAF <1%), a Hardy-Weinberg equilibrium (p<10^−6^) and missingness rates >5% were also excluded. Population stratification was checked by clustering the samples using principal component analysis. After the QC procedure, a total of 2,527 cognitively unimpaired individuals from the ALFA parent cohort were genetically characterized.

Imputation of genetic variants was carried out using the Michigan Imputation Server with the *Haplotype Reference Consortium* Panel (HRC r1.1 2016) (Das et al., 2016) following default parameters and established guidelines. Mismatched and monomorphic genetic variants were removed during pre-imputation filtering. Genetic variants with invalid alleles were switched and flipped in the GWAS sets (Human Reference Panel: GRCh37 hg19 b37 humanG1Kv37). Phasing was performed using the EAGLE software v.2.4, and imputation results were rechecked using standard Michigan imputation server QC parameters (imputation quality > 0.2 and MAF >1%).

### 2.3. Statistical Analysis

Differences in demographic characteristics were assessed according to *APOE*-ε*4* carriership, AT groups and sex, using chi-square tests for categorical variables and parametric (t-test, ANOVA) and non-parametric tests (Wilcox test or Kruskal-Wallis) for continuous normally and non-normally distributed variables, as appropriate. Pairwise comparison p-values were also provided, adjusted for false discovery rate correction.

Polygenic risk scores (PRS) were computed using PRSice version 2 (Choi & O’Reilly, 2019) to assess polygenicity. PRSice computes PRS by summing all SNP alleles carried by participants, weighting them by the SNP allele effect size estimated in a previous GWAS, and normalizing the score by the total number of SNPs included. PRS were calculated in representative genetic variants per linkage disequilibrium block (LD) (clumped variants), using a cut-off for LD of r^2^ > 0.1 in a 250-kb window. For PRS calculation of AD and AD CSF biomarkers, the *APOE* region was additionally excluded (chr19:45,409,011-45,412,650; GRCh37/hg19). Results were displayed at a restrictive threshold, 5x10^−6^. A total of 40 PRSs were calculated based on recently published GWAS, distributed in 6 main categories of diseases and conditions: AD Biomarkers (Jansen et al., 2022), neurodegenerative diseases (Wightman et al., 2021; Ferrari et al., 2014; Nalls et al., 2019), cerebrovascular diseases (Canela-Xandri et al., 2018; Malik et al., 2018), brain endophenotypes of AD (Smith et al., 2021; Persyn et al., 2020; Knol et al., 2020), AD risk factors (Meier et al., 2019; Okbay et al., 2016; Wray et al., 2018; Day et al., 2018; Yengo et al., 2018; Pulit et al., 2019; Teslovich et al., 2010; Canela-Xandri et al., 2018; Jansen et al., 2019), and aging biomarkers (Codd et al., 2021; Pilling et al., 2016; Atkins et al., 2021; Gibson et al., 2019). Further details on the PRSs can be found in **Supplementary Table 1**. In addition, individuals were classified into three groups based on the distribution of PRSs for AD, Aβ42 and pTau: low genetic predisposition (PRS ≤ percentile 10), intermediate genetic predisposition (percentile 10 < PRS ≤ percentile 90), and high genetic predisposition (PRS > percentile 90).

Additionally, significant differences in the median values (Wilcoxon test) and variances (Levene test) of the PRSs were evaluated between *APOE*-ε*4* carriers, AT groups in the ALFA+ cohort, and sex.

Finally, the AD predisposition in ALFA was compared (pairwise median test) to that across the full disease spectrum in the ADNI database by including controls (CN) (N=530), amyloid positive individuals with mild cognitive impairment (MCI) (N=598), and AD dementia patients (N=205). ALFA+ participants were further stratified according to Aβ42/40 status (A+, N=134; A-, N=246).

## 3. Results: Genetic characterization of the ALFA parent cohort

### 3.1. Demographic characteristics and *APOE* genotype distribution

A characteristic feature of the ALFA study is the high prevalence of *APOE-*ε*4* carriers compared with the general European population (35.6% *vs*. 14%; p□<□0.001) (Ward et al., 2012), and the high prevalence of homozygotes in the ALFA+ subsample [**Table 1, Figure 2**]. Significant differences in demographic characteristics were found between *APOE-*ε*4* non-carriers (N=1,624; 64.39%) and carriers (N=898; 35.61%). Non-carriers were on average older than ε*4*-homozygous individuals (p=0.024). The percentage of women in the non-carrier group was higher than in the *APOE*-ε*4* heterozygotes one (p=0.038). We also observed a higher percentage of individuals with a family history of AD in *APOE*-ε*4* heterozygotes compared to non-carriers (p<0.001) [**Figure 2A, Supplementary Table 2**]. In ALFA+, a total of 8% (N=31) of the individuals were *APOE*-ε*4* homozygous. Moreover, among *APOE*-ε*4* carriers, 8.2% of individuals were A+T+ (*vs*. 7.5% among non-carriers), 41.3% were A+T-(*vs*. 10.9% among non-carriers), and 50.5% were A-T-(*vs*. 81.6% among non-carriers) [**Figure 2B, Supplementary Table 3-4**]. Notably, there was a higher percentage of A+ individuals among *APOE-*ε*4* carriers compared with non-carriers (49.5% *vs*. 18.4%; p<0.001). Similar to the overall sample, the proportion of women was higher in the non-carrier group compared to the *APOE-*ε*4* heterozygotes one (p=0.029). No significant differences were found in the distribution of individuals according to family history of AD.

**Table 1.**
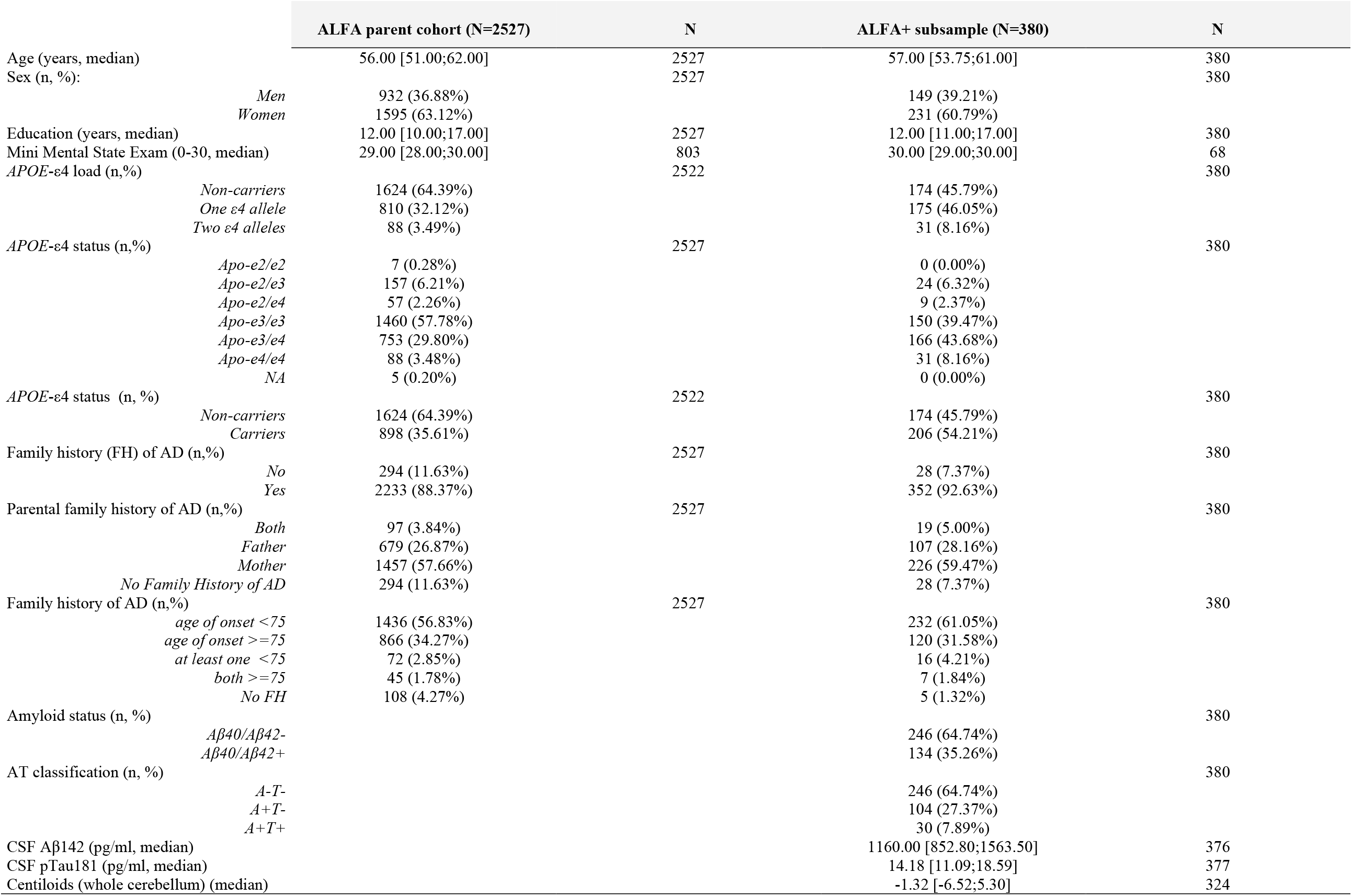
Demographic characteristics and APOE genotypes distribution in the ALFA parent cohort as well as in the ALFA+ subsample. A total of 5 individuals presented NA values in APOE-ε4 characterization. A total of 13 individuals with A-T+ profile were excluded from the ALFA+ subsample. Available CSF biomarkers data after outliers removal. A total of 56 individuals presented NA values in Centiloids.

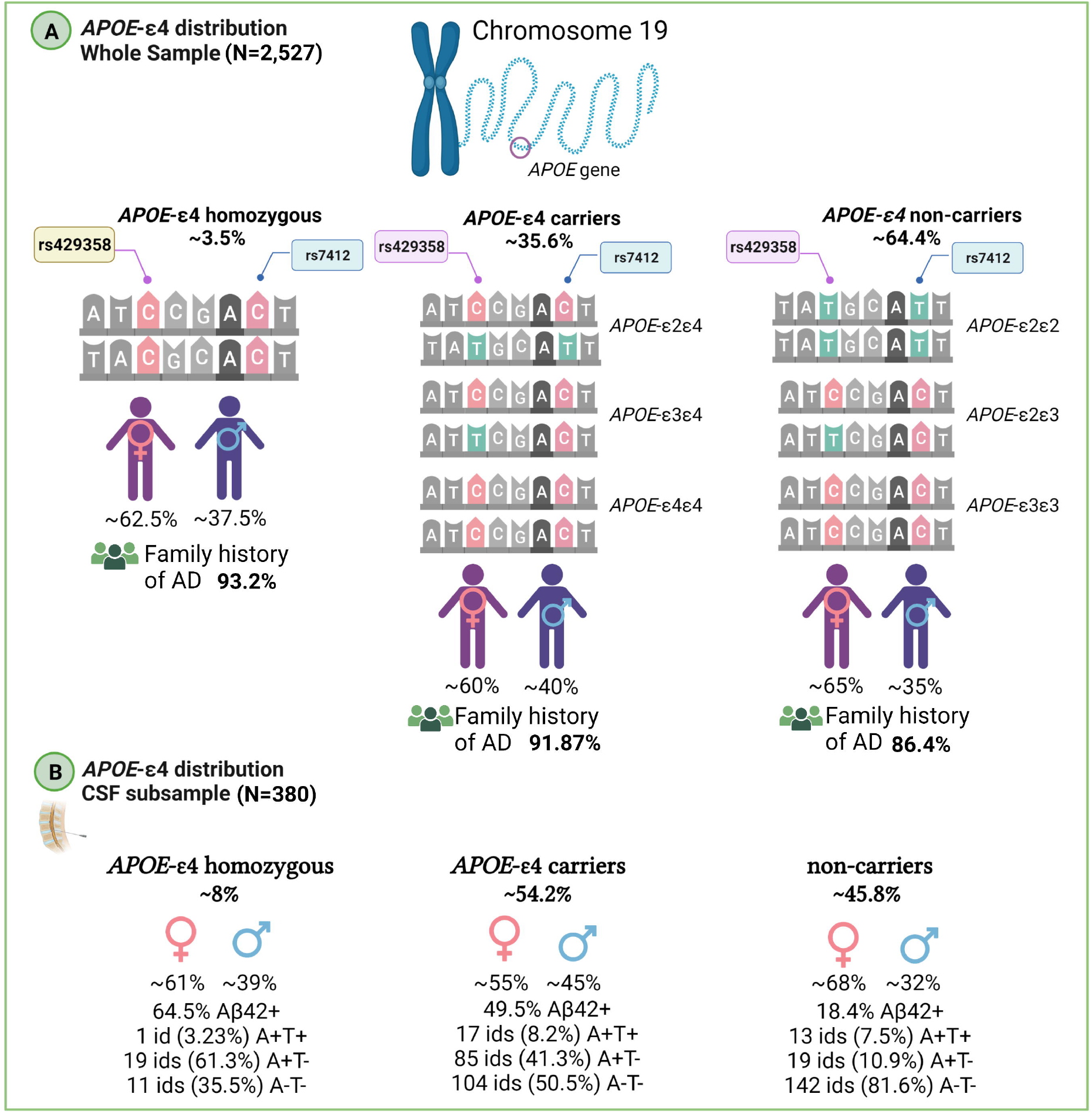

### 3.2. Polygenic characterization of ALFA participants

PRS of AD (PRS-AD) showed the highest variability in the sample (IQR=0.25) as well as frontotemporal dementia (FTD) subtypes (IQR>0.035), sleep risk factors and biological aging phenotypes (IQR>0.04), whereas Parkinson’s disease (IQR=3·10^−5^), cerebral infarction (IQR=8·10^−5^), and intracerebral hemorrhage (IQR=2·10^−5^) were the main conditions in which subjects had more homogeneous scores (IQR∼0) [**Figure 3, Supplementary Table 5**].

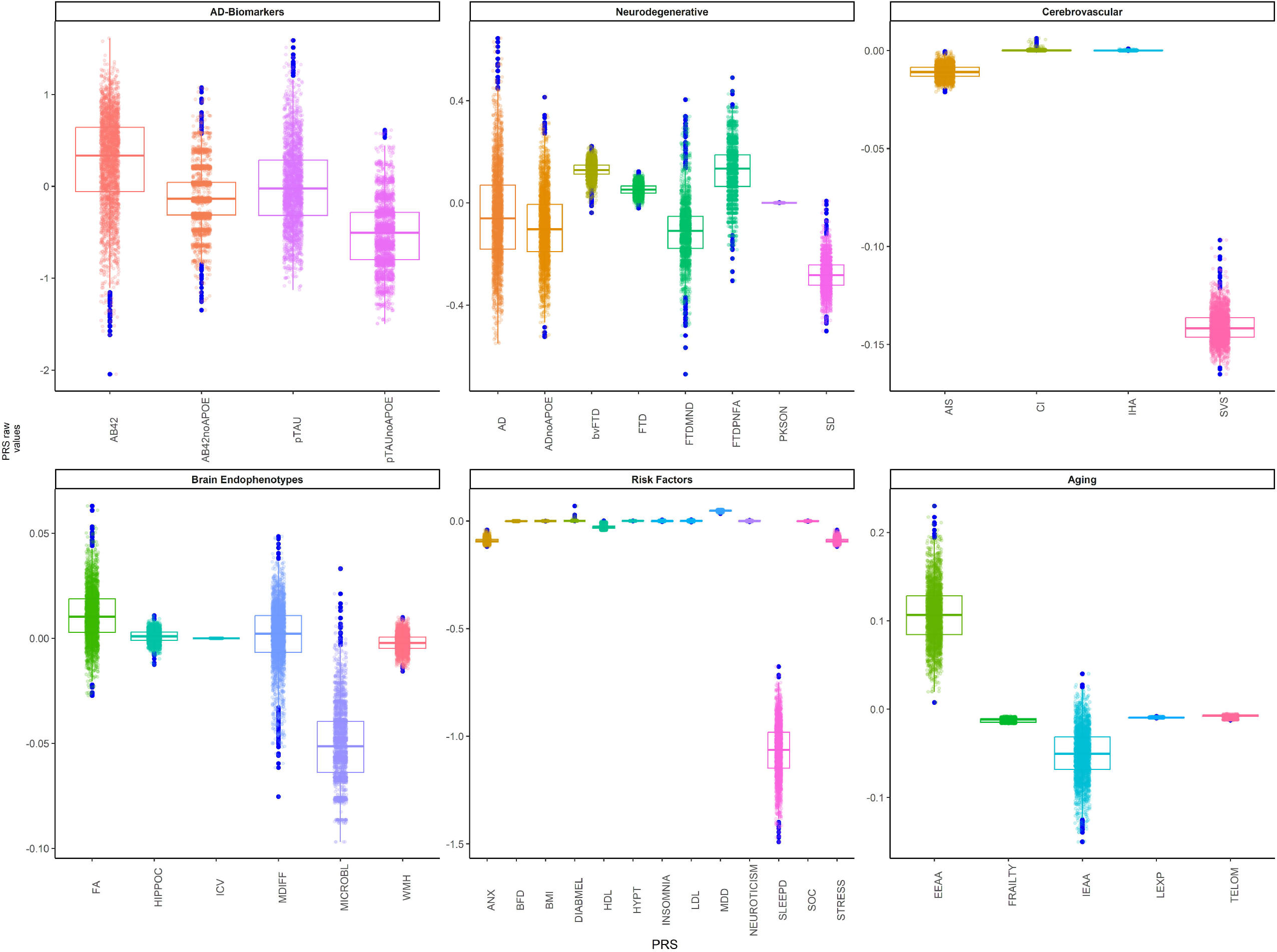

We observed that ε*4* homozygotes had higher scores for AD and FTD, AD biomarkers, and some brain endophenotypes, risk factors and aging conditions compared with heterozygotes and non-carriers [**Supplementary Figure 2**]. Moreover, we observed that A+T-individuals had higher scores for PRS-AD (p=0.006) as well as lower median PRS of CSF A[42 levels than A-T-(p∼0). In addition, A+T+ showed higher genetic predisposition to WMH than A-T-(p=0.006). Similarly, A+T-showed a higher median for the risk score of microbleeds and longer sleep duration than A-T-(p<0.018) [**Supplementary Figure 3, Supplementary Table 6**]. Finally, only significant differences were found between sex for genetic predisposition to social isolation and sleep duration [**Supplementary Figure 4, Supplemental Table 7**].

### 3.3. Characteristics of individuals with AD genetic predisposition

We found significant differences in the percentage of ε*4* carriers between individuals at high (81%), intermediate (33.3%), and low genetic predisposition to AD (7.94%) (p<0.001) [**Supplementary Table 8**]. However, when the *APOE* region was excluded from the PRS-AD, the proportion of ε*4* carriers and non-carriers was found to be balanced. For both, the proportion of individuals with a positive family history of AD was higher in the group with high genetic predisposition to AD than in the other groups. In ALFA+, we also observed a higher percentage of ε*4* carriers in individuals with high genetic predisposition to AD, and 21.2% were ε*4*-homozygous [**Supplementary Table 9**]. Additionally, the proportion of A+T- and A+T+ was also significantly higher in this group (p<0.05). We did not find any differences in the proportion of women and men across groups.

When we classified individuals according to their genetic predisposition to higher CSF-Aβ42 levels, we found that the proportion of *APOE*-ε*4* carriers and, specifically, homozygotes was significantly higher in the genetically predisposed group to present abnormal levels of CSF-Aβ42 (low genetic predisposition to higher CSF-Aβ42 levels) compared to the other groups (p<0.001). However, when we removed the *APOE* region from the PRS-Aβ42, we did not observe significant differences in the percentage of *APOE*-ε*4* carriers among groups [**Supplementary Table 10**]. Similarly, we found a higher proportion of *APOE-*ε*4* carriers, and specifically, homozygotes, in the group of individuals at higher genetic predisposition to display higher CSF-pTau levels (at risk group) (p<0.001). However, we did not find these differences when removing the effect of the genetic variants in the *APOE* region [**Supplementary Table 11**]. The above results were also assessed for the subgroup of individuals with available CSF biomarkers from the ALFA+ study. Significant differences were also found in the distribution of *APOE-*ε*4* carriers among genetic groups based on their genetic predisposition to CSF Aβ42 levels [**Supplementary Table 12**]. As in the whole sample, higher percentages of both *APOE*-ε*4* heterozygotes and homozygous were found in the group at lower genetic predisposition to higher CSF Aβ42 levels compared with the high and intermediate groups (p<0.001). Moreover, in this group we found a higher percentage of A+ individuals compared with the intermediate (p=0.008) and high (p=0.008) genetic groups. In the low genetic group, we found a higher percentage of A+T+ individuals than in the higher group (p=0.012) but lower than in the intermediate group (p=0.005). These differences were not observed when removing the *APOE* region. Significant differences were also observed between genetic groups based on their genetic predisposition to CSF pTau levels [**Supplementary Table 13**]. As in the whole sample, higher percentages of both *APOE-*ε*4* carriers and homozygous were found in the the group showing high genetic predisposition to higher CSF pTau levels compared with the low and intermediate groups (p<0.001). Moreover, in the high group, we found a higher percentage of A+ individuals as well as a higher percentage of A+T+ individuals compared with the low and intermediate genetic groups (pairwise comparisons p<0.05). Non-significant differences were observed after removing the *APOE* region.

### 3.4. AD genetic risk prediction in the whole disease spectrum

We observed that the median value of the PRS-AD increased along the AD *continuum* **[Figure 4]**. In ADNI, significant differences in the median score of PRS-AD were observed between CN and MCI (p<0.001) and between CN and AD (p<0.001). Significant differences were also found between MCI and AD subjects (p<0.05). When participants from the ALFA parent cohort were included in the spectrum, we found that the median value of the PRS-AD in ALFA participants was higher than in CN from ADNI (p<0.001) although was significantly lower than in MCI (p<0.01) and AD (p<0.001) **[Figure 4A]**. When ADNI and ALFA+ individuals were stratified by Aβ status, we found that the genetic predisposition was higher in ALFA+ A+ than in ALFA+ A-(p<0.001) [**Figure 4B]**. Moreover, the median PRS-AD in ALFA+ A+ individuals was higher compared both to CN A-(p<0.001) and CN A+ (p<0.05). Although there were no significant differences between them, ALFA+ A+ individuals showed a higher median value of the PRS-AD than MCI A+. Non-significant differences were found between ALFA+ A+ and AD individuals. [**Supplementary Table 15**].

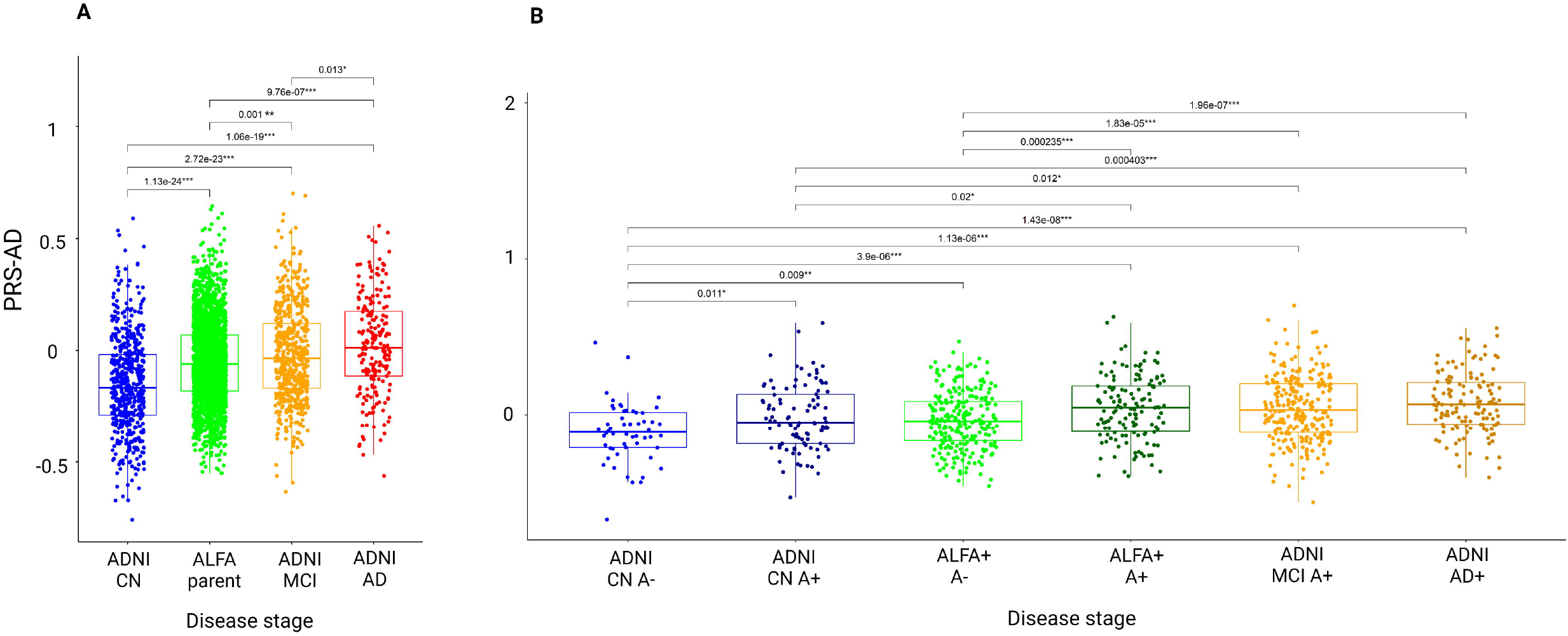

Comparisons stratifying by other risk factors were assessed **[Figure 5]**. We found significant differences among *APOE-*□*4* groups for all disease-stage groups [**Figure 5A**]. The higher the number of □*4* alleles, the higher the median value of the PRS-AD. Neither in ALFA A-nor in A+ significant differences were found in the median PRS-AD between □*4* allele heterozygous and homozygous. Nonetheless, in both groups, both □*4* allele heterozygous and homozygous displayed higher values for the median PRS-AD than non-carriers (p<0.001). Differences were also assessed stratifying by AT profiles [**Figure 5B**]. In the ALFA+ sample, A+T-individuals showed higher median PRS-AD than A-T- (p<0.001). In ADNI, CN that were A+T+ displayed higher median values than A-T- (p<0.01). In MCIs, significant differences were found between all pairwise comparisons (p<0.05), while in AD patients, non-significant differences were found in the median value of the PRS-AD among AT groups. Finally, we did not observe significant sex-differences within ADNI nor in ALFA groups [**Figure 5C**].

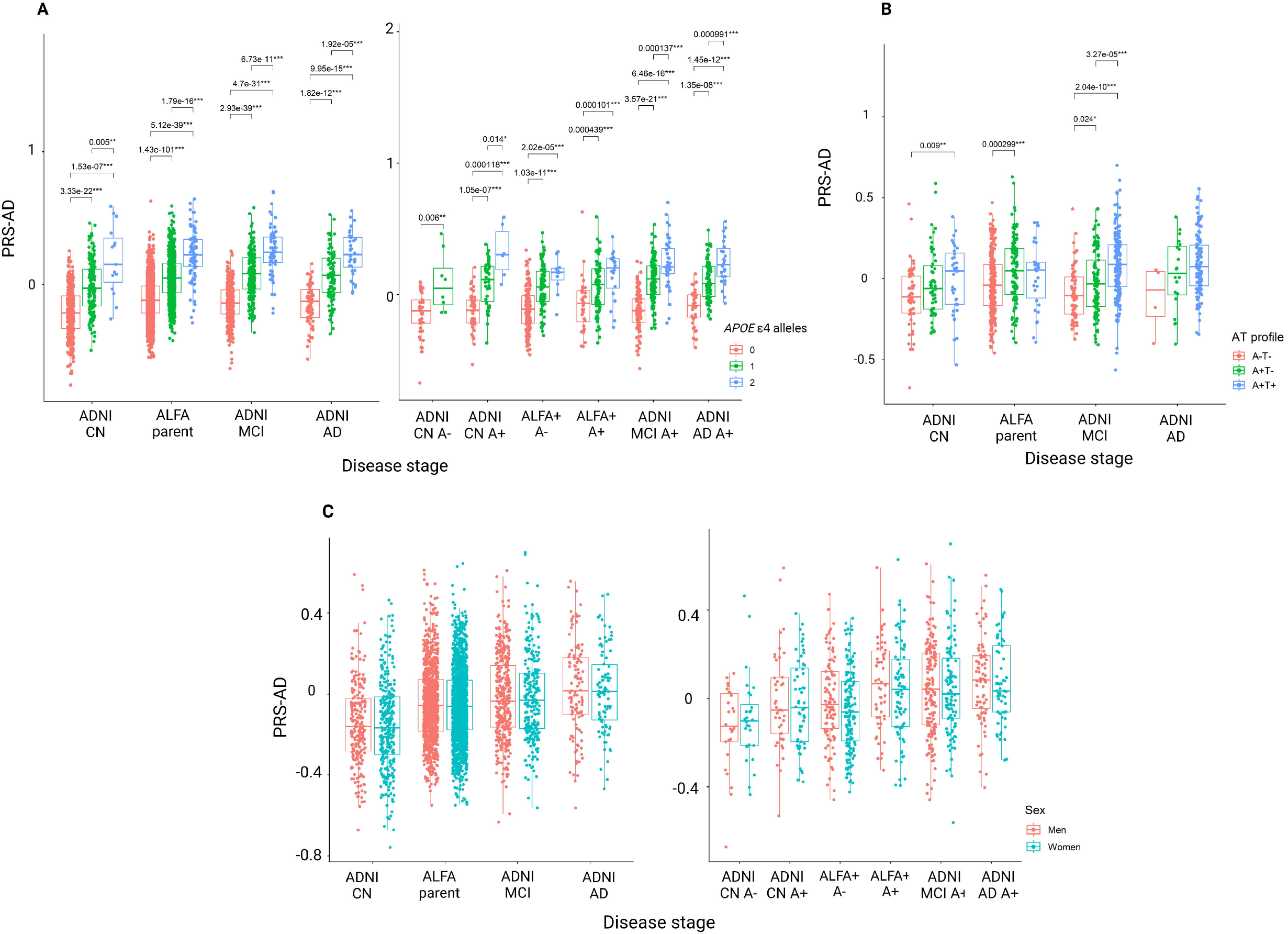

## 4. Discussion and future directions

The genetic characterization of ALFA parent cohort participants enabled us to investigate the genetic predisposition to neurodegenerative and other complex diseases in cognitively unimpaired individuals, many of whom were in the preclinical AD *continuum*. Our results showed that the ALFA parent cohort, and particularly ALFA+, are highly enriched for genetic risk factors for AD, making them suitable for studying early pathophysiological alterations in AD.

The study also revealed an increased genetic predisposition to AD in ALFA participants, even in the absence of the *APOE-*□*4* allele. This suggests that other variants (*APOE*-independent mechanisms) associated with AD appear to compensate for or complement the effect of *APOE* in genetic predisposition to AD, which is consistent with other studies (Luckett et al., 2022; Skoog et al., 2021). This illustrates how the use of PRS can reveal additional pathophysiological pathways with potential for prevention. (Daunt et al., 2021), which is relevant since the pathophysiology of AD has not yet been fully elucidated and the best biological characterization to date focuses on the determination of AD biomarkers (Milà-Alomà et al., 2020; Van Hulle et al., 2021). Indeed, individuals at genetic predisposition to display abnormal CSF Aβ42 levels were found to be enriched for *APOE-*ε*4* homozygosity, regardless of the inclusion of the *APOE* region, consistent with well-established findings reporting that the main effect of *APOE* in AD is linked to early alterations in Aβ42 metabolism (Salvadó, et al., 2021).

We also compared the PRS-AD with that in clinical groups in ADNI and found that ALFA+ participants, particularly those already on the AD *continuum* (A+), had a higher genetic risk profile than CN in ADNI, and similar to that of the clinical groups (MCI, AD). Overall, these results support that the ALFA population is well-suited to detect early pathogenic events in AD, both in *APOE*-associated and independent biological pathways. The study also confirmed that cognitively unimpaired individuals with abnormal AD biomarkers had a higher genetic predisposition to AD, as shown by the distribution of PRS-AD against AT groups. These results suggest that PRSs could be used as a proxy for risk stratification of unimpaired individuals (Vlachakis et al., 2021; Desikan et al., 2017). However, further research is needed to establish the additional information that PRSs provide in addition to biomarkers and their utility in research and clinical practice (Zettergren et al., 2021).

Moreover, individuals showing AD pathological changes (A+T-) showed higher genetic predisposition to AD, WMH, long sleep duration and microbleed counts compared to subjects with normal levels of AD biomarkers (A-T-). We did not detect any differences in PRS variability in T+ individuals, likely due to the low sample size of this group at the moment. These findings suggest different patterns of genetic predisposition in the Alzheimer’s *continuum* and/or increased susceptibility to comorbidities (Grande et al., 2021; Maciejewska et al., 2021). For instance, our results imply a possible common pathological mechanism linking genetic predisposition to cerebrovascular disease and sleep measurements to the development of amyloid-beta pathology at the earliest stages of the AD *continuum*. Further studies that consider scores regardless of the inclusion of the *APOE* region will additionally give insights into the biological interpretation of the results.

We observed slight differences in variability of genetic scores between sexes. Analyses using sex-specific summary statistics will help to better determine the mechanisms associated with such differences and to understand structural and functional differences in the brain, as well as cognitive and age-related processes (Deming et al., 2018; Dumitrescu et al., 2020). In addition, previous studies suggest the influence of sex-specific variants with small effects or more complex mechanisms involving epigenetic changes, gene-environment interactions, or genetic variants within sex chromosomes that should be incorporated into future studies (Silva et al., 2022; Wang et al., 2021).

The complex nature of AD, additionally leads to the use of other modalities for characterizing biological pathways involved in AD processes that can improve our understanding of how molecular differences relate to *in vivo* neuroimaging phenotypes (Arnatkeviciute et al., 2019). Moreover, techniques such as Mendelian Randomization can provide causal links between modifiable risk factors and AD pathology and its downstream consequences (Andrews et al., 2021) with potential impact to establish primary and secondary (preclinical stage) prevention strategies (Rodríguez-Fernández et al., 2022).

Finally, collaborating with global initiatives like the UNITED (Uncovering Neurodegenerative Insights Through Ethnic Diversity) Consortium (Adams et al., 2019), the European Prevention of Alzheimer’s Dementia (EPAD) cohort (Ritchie et al., 2020), and the Quantitative amyloid PET in Alzheimer’s disease (AMYPAD) initiative (Lopes Alves et al., 2020), and using publicly available genetic data from other resources are essential (Bycroft et al., 2018) will be crucial to better unravel the genetic mechanisms underlying AD worldwide taking also into account diverse populations.

## Data Availability

All data produced in the present study are available upon reasonable request to the authors.

## Conflict of interest statement

JLM is currently a full-time employee of Lundbeck and has previously served as a consultant or at advisory boards, or has given lectures in symposia sponsored by the following for-profit companies: Roche Diagnostics, Genentech, Novartis, Lundbeck, Oryzon, Biogen, Lilly, Janssen, Green Valley, MSD, Eisai, Alector, BioCross, GE Healthcare, ProMIS Neurosciences. JDG has received speaker’s or consultant’s fees from Philips Nederlands, Roche Diagnostics and Biogen and research support from GE Healthcare, Roche Diagnostics and Hoffmann-La Roche. MSC has served as a consultant and at advisory boards for Roche Diagnostics International Ltd, has given lectures in symposia sponsored by Roche Diagnostics, S.L.U, Roche Farma, S.A and Roche Sistemas de Diagnósticos, Sociedade Unipessoal, Lda. and research support from Roche Diagnostics International Ltd. GSB has served as a consultant for Roche Farma, S.A. The remaining authors declare that they have no conflict of interest.

## Funding

The project leading to these results has received funding from “la Caixa” Foundation (ID 100010434), under agreement LCF/PR/GN17/50300004, the Health Department of the Catalan Government (Health Research and Innovation Strategic Plan (PERIS) 2016-2020 grant# SLT002/16/00201) and the Alzheimer’s Association (Grant AARG-19-618265). Additional support has been received from the Universities and Research Secretariat, Ministry of Business and Knowledge of the Catalan Government under grant no. 2021_SGR_00913. All CRG authors acknowledge the support of the Spanish Ministry of Science, Innovation, and Universities to the EMBL partnership, the Centro de Excelencia Severo Ochoa, and the CERCA Programme / Generalitat de Catalunya. NV-T and OG-R receive funding from the MCIN/AEI/10.13039/501100011033 and the European Union NextGenerationEU/PRTR, through the Juan de la Cierva Incorporación Programme (IJC2020-043216-I and IJC2020-043417-I respectively). MS-C receives the support of a fellowship from “la Caixa” Foundation (ID 100010434) and from the European Union’s Horizon 2020 research and innovation programme under the Marie Skłodowska-Curie grant agreement No 847648. The fellowship code is LCF/BQ/PR21/11840004.

## Standard Protocol Approvals, Registrations, and Patient Consents

The study was conducted in accordance with the directives of the Spanish Law 14/2007, of 3rd of July, on Biomedical Research (Ley 14/2007 de Investigación Biomédica). The ALFA study protocol was approved by the Independent Ethics Committee Parc de Salut Mar Barcelona and registered at Clinicaltrials.gov (Identifier: <u>NCT01835717</u>). All participants accepted the study procedures by signing the study’s informed consent form that had also been approved by the same IRB.

## Acknowledgments

This publication is part of the ALFA study. The authors would like to express their most sincere gratitude to the ALFA project participants and relatives without whom this research would have not been possible. Collaborators of the ALFA study are: Müge Akinci, Federica Anastasi, Annabella Beteta, Raffaele Cacciaglia, Lidia Canals, Alba Cañas, Carme Deulofeu, Maria Emilio, Irene Cumplido-Mayoral, Marta del Campo, Carme Deulofeu, Ruth Dominguez, Maria Emilio, Sherezade Fuentes, Marina García, Laura Hernández, Gema Huesa, Jordi Huguet, Laura Iglesias, Esther Jiménez, David López-Martos, Paula Marne, Tania Menchón, Paula Ortiz-Romero, Eleni Palpatzis, Wiesje Pelkmans, Albina Polo, Sandra Pradas, Mahnaz Shekari, Lluís Solsona, Anna Soteras, Laura Stankeviciute, Núria Tort-Colet and Marc Vilanova.

We would like to additionally express our sincere gratitude to Prof. Kaj Blennow and Prof. Henrik Zetterberg for their invaluable assistance in acquiring the biomarkers used in the ALFA+ study. We would also like to acknowledge the guidance and expertise of Prof. Roderic Guigó in the acquisition and modellization of the genetic data, and Prof. Anna González-Neira for her guidance in genetic quality control. Finally, we extend our heartfelt appreciation to Prof. Jordi Camí for his significant contribution to the design of the ALFA parent cohort. The authors thank Roche Diagnostics International Ltd for providing the kits to measure CSF biomarkers, and the laboratory technicians at the Clinical Neurochemistry Lab in Mölndal, Sweden, who performed the analyses. COBAS, COBAS E, and ELECSYS are trademarks of Roche.

Data collection and sharing for this project was funded by the Alzheimer’s Disease Neuroimaging Initiative (ADNI) (National Institutes of Health Grant U01 AG024904) and DOD ADNI (Department of Defense award number W81XWH-12-2-0012). ADNI is funded by the National Institute on Aging, the National Institute of Biomedical Imaging and Bioengineering, and through generous contributions from the following: AbbVie, Alzheimer’s Association; Alzheimer’s Drug Discovery Foundation; Araclon Biotech; BioClinica, Inc.; Biogen; Bristol-Myers Squibb Company; CereSpir, Inc.; Cogstate; Eisai Inc.; Elan Pharmaceuticals, Inc.; Eli Lilly and Company; EuroImmun; F. Hoffmann-La Roche Ltd and its affiliated company Genentech, Inc.; Fujirebio; GE Healthcare; IXICO Ltd.; Janssen Alzheimer Immunotherapy Research & Development, LLC.; Johnson & Johnson Pharmaceutical Research & Development LLC.; Lumosity; Lundbeck; Merck & Co., Inc.; Meso Scale Diagnostics, LLC.; NeuroRx Research; Neurotrack Technologies; Novartis Pharmaceuticals Corporation; Pfizer Inc.; Piramal Imaging; Servier; Takeda Pharmaceutical Company; and Transition Therapeutics. The Canadian Institutes of Health Research is providing funds to support ADNI clinical sites in Canada. Private sector contributions are facilitated by the Foundation for the National Institutes of Health (www.fnih.org). The grantee organization is the Northern California Institute for Research and Education, and the study is coordinated by the Alzheimer’s Therapeutic Research Institute at the University of Southern California. ADNI data are disseminated by the Laboratory for Neuro Imaging at the University of Southern California.

We thank all the Consortium members involved in the analysis and generation of summary statistics used in this project.

CSF Amyloid Aβ42 (https://www.ebi.ac.uk/gwas/ Accession ID GCST90129599); CSF Tau phosphorylated (https://www.ebi.ac.uk/gwas/ Accession ID GCST90129600); Leukocyte Telomere Length (Requested to corresponding Author); Fractional Anisotropy (http://www.kp4cd.org/dataset_downloads/stroke); Hippocampal volume (http://ftp.ebi.ac.uk/pub/databases/gwas/summary_statistics/GCST90002001-GCST90003000/GCST90002711/); Mean Diffusivity (http://www.kp4cd.org/dataset_downloads/stroke); Microbleeds (http://www.kp4cd.org/dataset_downloads/stroke); White Matter Hyperintensities (http://www.kp4cd.org/dataset_downloads/stroke); Ischemic Stroke (Requested to corresponding Author); Cerebral infarction (http://geneatlas.roslin.ed.ac.uk); Small Vessel Disease (Requested to corresponding Author); Alzheimer’s Disease (https://ctg.cncr.nl/software/summary_statistics); Frontotemporal Dementia and subtypes (https://ifgcsite.wordpress.com/data-access/); Parkinson’s Disease (https://drive.google.com/file/d/1FZ9UL99LAqyWnyNBxxlx6qOUlfAnublN/view?usp=sharing); Semantic Dementia (https://ifgcsite.wordpress.com/data-access/); Anxiety Disorder (https://ipsych.dk/en/research/downloads/); Body Mass Index (https://portals.broadinstitute.org/collaboration/giant/index.php/GIANT_consortium_data_files); HDL Cholesterol (http://csg.sph.umich.edu/willer/public/lipids2010/); Hyperthyroidism/thyrotoxicosis (http://geneatlas.roslin.ed.ac.uk); Hypothyroidism/myxoedema (http://geneatlas.roslin.ed.ac.uk); Hypertension (http://geneatlas.roslin.ed.ac.uk); Insomnia (https://ctg.cncr.nl/software/summary_statistics); LDL Cholesterol (http://csg.sph.umich.edu/willer/public/lipids2010/); Life Expectancy (http://ftp.ebi.ac.uk/pub/databases/gwas/summary_statistics/GCST003001-GCST004000/GCST003395/results.UKBiobank_9millionSNPs.parents_lifespan.Pilling_et_al_2016_top_1_percent.txt); Sleep Duration (https://ctg.cncr.nl/software/summary_statistics); Stress Disorder (https://ipsych.dk/en/research/downloads/). Further details can be found in Supplementary Table 1. Schematic representations were created with Biorender.com.

## Notes

### Author Declarations

Standard Protocol Approvals, Registrations, and Patient Consents The ALFA parent cohort (Identifier: NCT01835717) and the ALFA+ study (ALFA-FPM-0311) were approved by the Independent Ethics Committee Parc de Salut Mar, Barcelona, and registered at ClinicalTrials.gov (identifier: NCT02485730). All participants signed the study's informed consent form that had also been approved by the Independent Ethics Committee, Parc de Salut Mar, Barcelona.

