## Supplementary figures and images for "Genetic characterization of the ALFA study: Uncovering genetic profiles in the Alzheimer’s *continuum*"

### Còpia de FS1_new.png

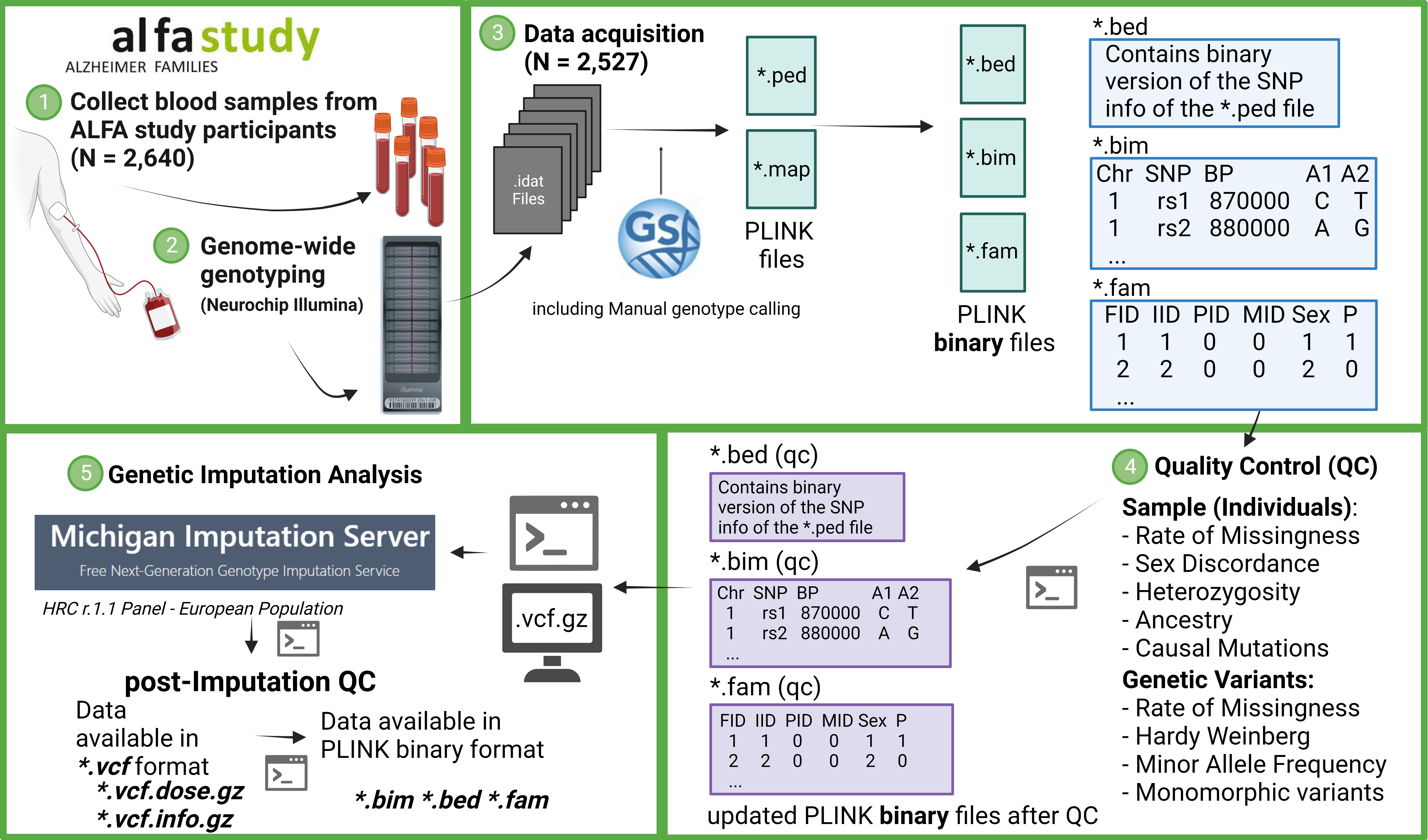

### Còpia de FS2_new.png

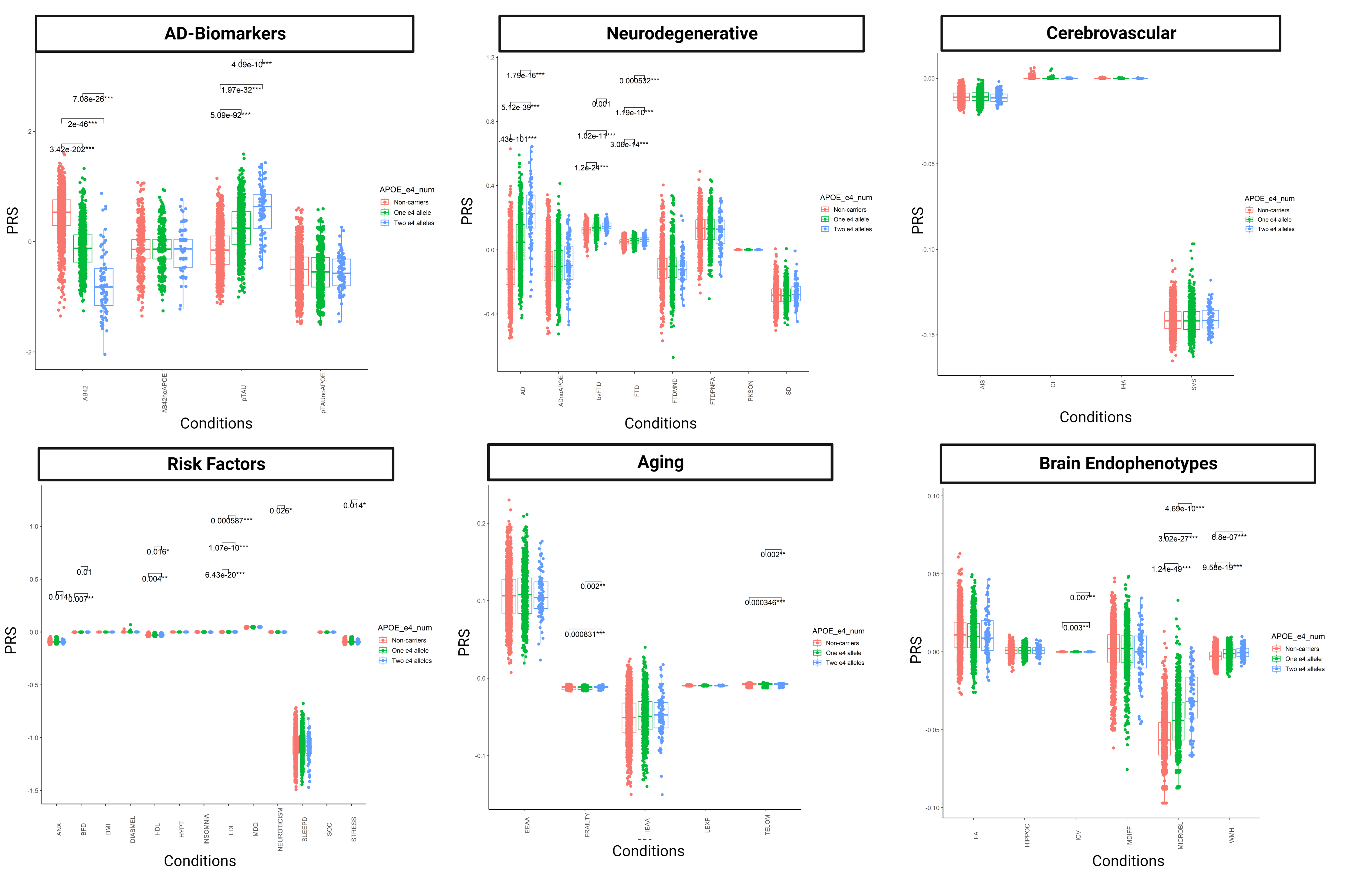

### Còpia de FS3_new.png

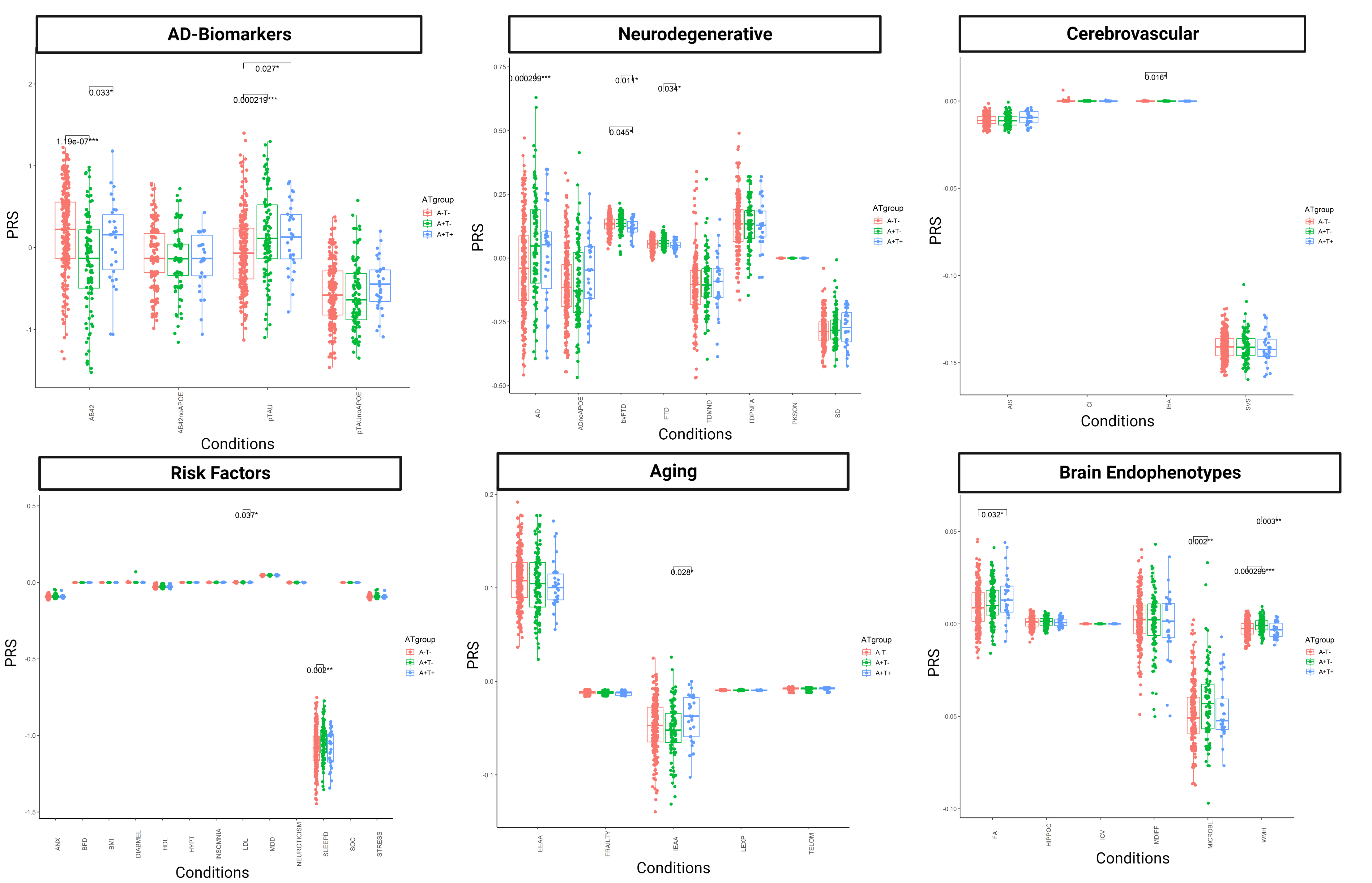

### Còpia de FS4_new.png

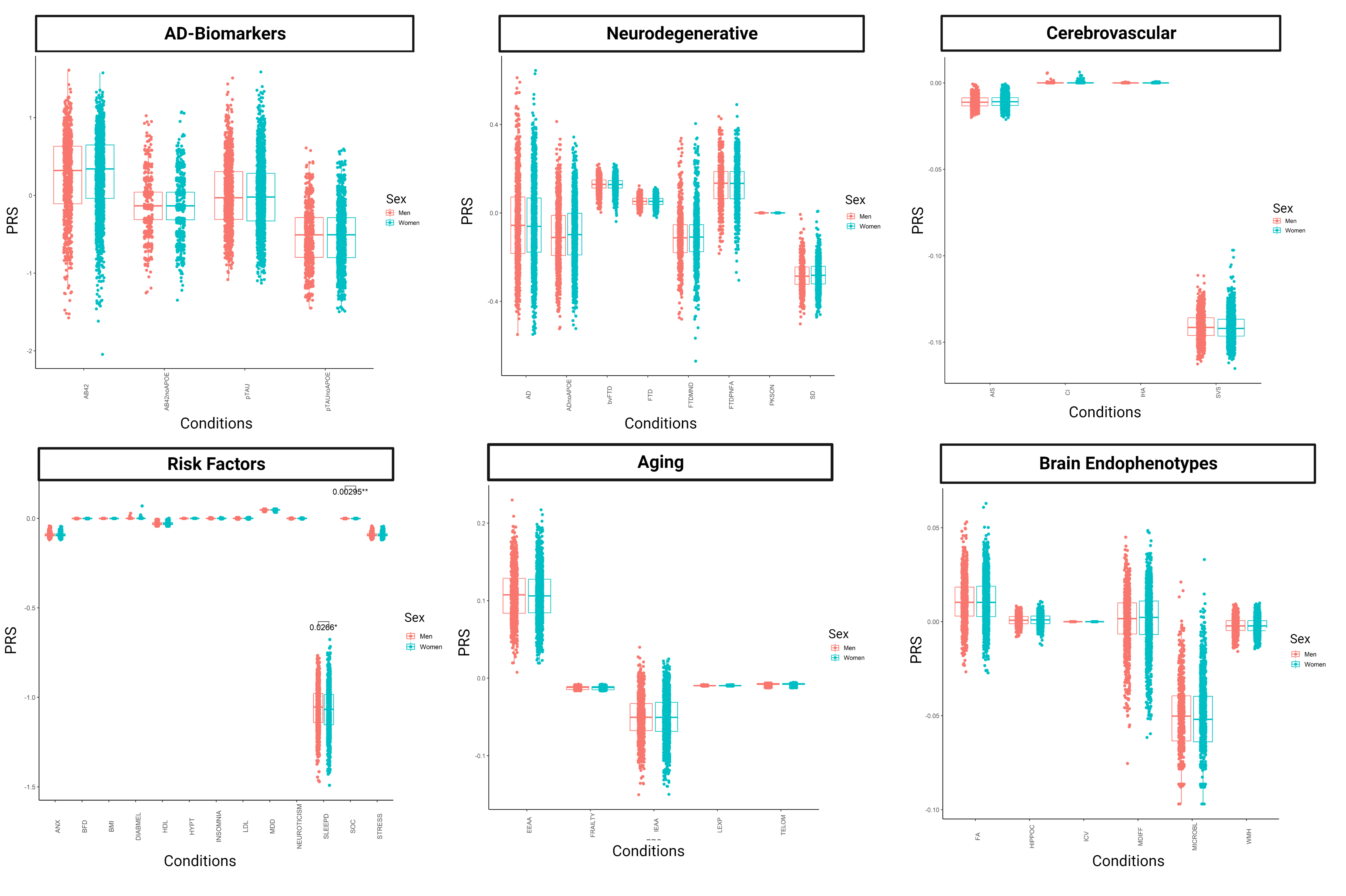
