## Supplemental Methods for "Genetic characterization of the ALFA study: Uncovering genetic profiles in the Alzheimer’s *continuum*"

**DNA extraction, Genotyping and data availability in ALFA**

DNA samples were obtained from whole blood samples by applying a salting out protocol. DNA was eluted in 800µl of H_2_O (Milli-Q®) and quantified using Quant-iTT PicoGreen® dsDNA Assay Kit (Life Technologies). The integrity of DNA was checked in a subset of samples by running a 1% agarose gel. All the samples were within specification. The DNA concentration for each sample was additionally normalized.

Genome-wide genotyping was performed in three different batches using the Illumina Infinium Neuro Consortium (NeuroChip) Array (build GRCh37/hg19) version 1.0 (batch 1: N = 936; batch 2: N = 1,080) and version 1.2 (batch 3: N = 624). An additional fourth batch (N = 297) (version 1.2) was performed including individuals who did not pass genetic quality control in previous batches, or for which blood samples were acquired *a posteriori*. The NeuroChip Array (Blauwendraat et al., 2017) contains a backbone consisting of ~310K tagging variants (Infinium HumanCore-24 v1.0 and Infinium HumanCore-24 v1.2) that densely cover ancestry informative markers for the determination of identity by descent and X chromosome single nucleotide polymorphisms (SNPs) for genetically sex determination. Additionally, NeuroChip Array contains a custom content of ~180K neurodegenerative disease-related variants of Alzheimer’s disease, Parkinson’s disease, Frontotemporal dementia, Amyotrophic Lateral Sclerosis, and other types of dementia.

Genotype calling was performed with the Illumina GenomeStudio 2.0 software. First, genotype clustering was done using the GenTrain 3.0 algorithm. We discarded the worst-performing samples by call rate (proportion of non-failed probes). For the present procedure, three call rate thresholds were tested (0.90, 0.95, 0.99). Then, we repeated the genotype clustering with GenTrain 3.0 using only the non-excluded samples. The genetic variants statistics for the samples were collected from each iteration and the best iteration was selected. Finally, we performed a final genotype calling using the pre-computed parameters.

**Extended Sample Characteristics**

*Cognitive outcomes*

Participants’ cognitive reserve was assessed with the administration of the Cognitive Reserve Questionnaire (Rami et al., 2011) consisting of 8 items, namely formal education, parental formal education, attendance to courses, occupation, musical education, languages spoken, frequency of reading, and cognitively stimulating activities. After an initial evaluation (for the specific inclusion criteria please refer to Molinuevo et al., 2016), a total of 2,527 eligible subjects were administered an experimental cognitive test battery for the potential detection of early impairment in longitudinal follow-ups. This battery assessed episodic verbal memory the Memory Binding Test [(Buschke et al., 2017; Gramunt et al., 2015)], psychomotor speed, visual processing, executive function, and non-verbal and verbal reasoning (Coding, Visual Puzzles, Digit Span, Matrix Reasoning, and Similarities of the WAIS-IV (Wechsler, 2012).

*Cognitive performance falling*

Outside the established cutoffs: Mini-Mental State Examination (MMSE) < 26, or Memory Impairment Screen (MIS) <6, or Time-Orientation subtest of the Barcelona Test II(TO-BTII) <68, or semantic fluency (animals; SF) <12. Clinical Dementia Rating scale (CDR)>0.

*Biomarkers*

A subset of the ALFA study participants were invited to take part in a nested longitudinal long-term study, referred to as ALFA+, in which more detailed phenotyping is performed. It entails the acquisition of both fluid (CSF, blood) and imaging (MRI and PET) biomarkers, as well as an extended cognitive assessment. Specifically, 419 individuals have been included and undergone ALFA+ baseline visit (October 2016 - December 2019), with the first follow-up visit currently ongoing (November 2019 - (expected) Q2 2022). Of them, 393 with available CSF and genetic data, and 286 with available PET and genetic data are included in the ALFA study. CSF samples were obtained by lumbar puncture following standard procedures (Milà-Alomà et al., 2020; Teunissen et al., 2014), and all the measurements were conducted at the Clinical Neurochemistry Laboratory, Sahlgrenska University Hospital, Mölndal, Sweden. Aβ pathology positivity (A+) and tau pathology positivity (T+) were defined by CSF Aβ42/40 ratio and CSF Mid(M)-p-tau181, respectively (Jack et al., 2016). The derived cut-offs are explained in detail in (Milà-Alomà et al., 2020) and were used to define AT groups (i.e. A+T+. A+T-, A-T-). Further details regarding CSF collection and analysis can be found in (Milà-Alomà et al., 2021). [^18^F]Flutemetamol (Aβ) and [^18^F]fluorodeoxyglucose (FDG) PET scans were acquired following a cranial computerized tomography (CT) scan for attenuation correction on a Biograph mCT scanner (Siemens Healthcare, Erlangen, Germany) at Hospital Clínic, Barcelona (Salvadó et al., 2019).

*Magnetic Resonance Imaging*

In addition, a subset of 1,426 participants with no contraindications to undergo MRI, were invited around 3 years later (June 2016 - August 2019) to a further visit that included a revision of their clinical, cognitive, and risk factor status together with the performance of an MRI. Available imaging sequences for most of the participants are isotropic high-resolution 3D T1-weighted imaging, high-resolution 3D T2-weighted imaging, ultrahigh-resolution multimodal inversion recovery, isotropic resting-state functional MRI, diffusion tensor imaging, multi-compartmental diffusion-weighted imaging, 3D T2-Fluid-attenuated inversion recovery, susceptibility-weighted imaging, arterial spin labeling, magnetic resonance spectrometry, multi-compartmental diffusion-weighted imaging, and cerebrospinal fluid flow imaging among others. All acquired imaging data were automatically transferred to and stored using the eXtensible Neuroimaging Archive Toolkit, XNAT (Marcus et al., 2007) framework which is accessible at barcelonabrainimaging.org (Huguet et al., 2021).

*Telomere length determination*

A total of 1,660 participants were selected for telomere length (TL) determinations based on the availability of biological samples (already stored at the biobank), cognitive, and MRI (see above) data (available at in-house databases). Samples were sent to the Harvard Cancer Center Genotyping & Genetics for Population Sciences Facility for TL determination using a high throughput version of the quantitative real-time polymerase chain reaction (qPCR)-based telomere assay. TL was measured in a single batch for all samples. TL was determined by real-time qPCR from the DNA extracted from peripheral blood leukocytes. First, DNA was quantified and normalized, then the relative TL was determined by a high-performance version of the real-time qPCR for telomeres. The assay was run on the Applied Biosystems 7900HT Sequence Detection System (Foster City, CA, USA). Laboratory personnel were blinded to participants’ characteristics, and all assays were processed in triplicate by the same technician and under identical conditions. Forty-five samples failed the assay, leaving reliable data for a total of 1,615 participants. Of them, 1,600 have available genetic data.

*Environmental Exposures*

We obtained environmental variables including levels of air pollution, indicators of green and blue spaces exposure, as well as diurnal and nocturnal noise levels. Individual levels of air pollution exposure included nitrogen oxides (NO_x_) and particulate matter (PM_2.5_ [particulate matter with aerodynamic diameter less than 2.5 μm], PM_10_ [less than 10 μm], PM coarse [PM_2.5–10_, i.e., coarse particulate matter, between 2.5 μm and 10 μm], and PM_2.5_ absorbance [PM_2.5_ light absorption]). Land Use Regression (LUR) models based on geographical information systems (GIS) were used to estimate individual levels of exposure to these air pollutants at the participant’s residential addresses (period 2009-2014) (Vert et al., 2017). Moreover, for each participant residential green and blue exposure indicators [surrounding greenness (NDVI), amount of green (land-cover), and access to major green spaces and blue spaces (forest, urban green area, agriculture area)] were generated for different buffers (100 m, 300 m and 500 m) (Gascon et al., 2018). Finally, participants’ residential noise daily and night levels were also measured in two different periods (2007 and 2012). A total of 2,425 participants with available genetic data are available in the study.
